# National Trends and Disparities in Hospitalization for Hypertensive Emergencies Among Medicare Beneficiaries, 1999–2019

**DOI:** 10.1101/2021.06.16.21259053

**Authors:** Yuan Lu, Yun Wang, Erica S. Spatz, Oyere Onuma, Khurram Nasir, Fatima Rodriguez, Karol E. Watson, Harlan M. Krumholz

## Abstract

**Importance:** In the last two decades, hypertension control in the U.S. population has not improved, and there are widening disparities. Less is known, however, about progress in reducing hospitalizations related to hypertensive emergencies.

**Objectives:** To describe trends in national hospitalization rates for hypertensive emergencies, overall and by demographic and geographical subgroups.

**Design, Setting and, Participants:** Serial cross-sectional analysis of Medicare fee-for-service beneficiaries aged 65 years or older between 1999 and 2019 using Medicare denominator and inpatient files.

**Main Outcome and Measures:** Trends in hospitalization for hypertensive emergencies, overall and by specific subgroups.

**Results:** The sample consisted of 397,238 individual Medicare fee-for-service beneficiaries. From 1999 through 2019, the annual hospitalization rates for hypertensive emergencies increased significantly from 51.5 to 125.9 per 100,000 beneficiary-years; this increase was most pronounced among the following subgroups: adults ≥85 years (66.8 to 274.1), females (64.9 to 160.1), Blacks (144.4 to 369.5), and Medicare-Medicaid insured (dual eligible, 93.1 to 270.0). Across all subgroups, Black adults had the highest hospitalization rate in 2019, and there was a significant increase in the differences in hospitalizations between Blacks and Whites from 1999 to 2019. Marked geographic variation was also present, with the highest hospitalization rates in the South (so-called “Stroke Belt”). Among 3,143 counties and county-equivalents included in the study, less than 1% of counties either had no change (n=7) or decreased (n=20) hospitalization rates since 1999. Among patients hospitalized for a hypertensive emergency, the observed 30-day all-cause mortality rate decreased from 2.6% to 1.7% and 30-day all-cause readmission rate decreased from 15.7% to 11.8%.

**Conclusions and Relevance:** Among Medicare fee-for-service beneficiaries aged 65 years or older, hospitalization rates for hypertensive emergencies increased substantially and significantly from 1999 to 2019. Black adults had the largest increase in hospitalization rates across age, sex, race, and dual-eligible strata. There was significant national variation, with the highest rates generally in the South.

**KEY POINTS:** *Question:* How have hospitalization rate for hypertensive emergencies among US adults aged 65 years and older changed between 1999 and 2019 and are there any differences across demographic and geographical subgroups?

*Findings:* In this serial cross-sectional study that included 397,238 individual Medicare fee-for-service beneficiaries, there was a marked increase in hospitalization rates for hypertensive emergencies from 1999 to 2019, and this increase was most pronounced among Black adults across age, sex, race, and dual-eligible strata. Significant national variation was observed, with the highest hospitalization rates generally in the South.

*Meaning:* Between 1999 and 2019, hospitalization rates for hypertensive emergencies increased substantially and differences across demographic and geographic subgroups persisted.

## BACKGROUND

In recent decades, the United States (US) has made little progress in hypertension control. Despite the availability of effective treatment and increase in hypertension diagnosis and awareness, national hypertension control (defined as blood pressure <140/90mmHg) has dropped from 54% in 2013-2014 to 44% in 2017-2018.^1^ Given the growing prevalence and devastating effects of uncontrolled hypertension, the Surgeon General recently issued a call to action to make hypertension control a national priority.^2^ A salient question is whether we have made any progress in preventing hospitalizations for hypertensive emergencies – acute severe blood pressure elevations that are associated with target organ damage and require urgent interventions.^3^ It is also unknown whether there are differences in hospitalization trends by age, sex, race and region over time.

Prior reports using data from the Nationwide Inpatient Sample indicate that hospitalization rates for hypertensive emergencies increased in the past decade;^4-6^ however, these studies focused on short time frames, were not able to capture the entire nation’s experience, did not assess demographic or geographic differences, and were not well positioned to assess trends due to sampling issues.^7^ Thus, we lack a contemporary, comprehensive, national perspective on hospitalization trends for hypertensive emergencies and associated outcomes, including readmission and mortality, and how they vary by demographic subgroups and geography.

The Centers for Medicare & Medicaid Services (CMS) is ideally positioned to provide information on trends in hospitalizations, mortality, and readmission outcomes nationally and by county. Accordingly, we studied all Medicare fee-for-service beneficiaries between 1999 and 2019 and evaluated hospitalization rates for hypertensive emergencies and longer-term outcomes, including 30-day readmission and 30-day mortality. Because substantial demographic and regional variation may exist in hypertension prevalence and outcomes, we evaluated rates of hospitalization and associated outcomes by demographic subgroups and county.

## METHODS

### Study Population

We identified all Medicare beneficiaries 65 years of age or older enrolled in the fee-for-service plan for at least 1 month from January 1999 to December 2019 using Medicare denominator files. For each year, we counted the total number of beneficiaries and calculated beneficiary-years to account for new enrollment, disenrollment, or death during the study period. We then linked beneficiary data with Medicare inpatient claims data to identify beneficiaries admitted with a primary discharge diagnosis of hypertensive emergency, hypertension urgency, or hypertension crisis (International Classification of Diseases, Ninth Revision, Clinical Modification codes [ICD-9 CM] of 401.0, 402.0x, 403.0x, 404.0x, and 405.0x from January 1, 1999, to September 31, 2015, ICD-10 CM code of I100, I119, I110, I120, I132, I150, and I158 from October 1, 2015, to September 31, 2016, ICD-10 CM code of I16.0, I16.1, I16.9 from October 1, 2016, to December 31, 2019, **Supplemental eTable 1**). No marked change in hospitalization was observed when switching codes from ICD-9 to ICD-10. Nevertheless, we reported the rates of hospitalization based on a 3-year interval to minimize the effect of changes in coding over time. We excluded hospitalizations with a secondary discharge diagnosis of hypertensive emergency since severe hypertension can occur in response to an acute illness and may not have been the primary driver of hospitalization. We obtained mortality data from the Medicare denominator files.

### Patient Characteristics

We determined the age, sex, and race (White, Black, Other) of beneficiaries and counted the number eligible for Medicaid for at least 1 month (dual eligible) for the Medicare fee-for-service beneficiaries who were hospitalized for hypertensive emergencies. The race variable was extracted from the Medicare enrollment database (EDB) and originates from Social Security Administration records. Before 1980, the Social Security Administration collected voluntary race data using the categories: White, Black, Other, and Unknown. The race of the beneficiary was assigned to the spouse. Given the EDB race variable is known to undercount Hispanics and other racial subgroups, analyses using race/ethnicity data from the EDB are generally restricted to the identification of differences between black and white patient populations.^8,9^ We ascertained comorbidities from secondary diagnosis codes as well as from principal and secondary diagnosis codes from all hospitalizations for 12 months before the index hospitalization; data from 1998 were used for hospitalizations in 1999. These comorbidities were classified using the Hierarchical Condition Categories method.^10,11^

### Outcomes

Our primary outcome was hospitalization for hypertensive emergencies. Secondary outcomes were 30-day all-cause mortality, 30-day all-cause readmission, and 30-day cause-specific readmission rates. We calculated hypertension-specific hospitalization rates by dividing the total numbers of hypertensive emergency discharges in each year by the corresponding person years of fee-for-service beneficiaries for that year.^12^ To standardize the follow-up period, we used 30-day mortality rate, defined as the rate of deaths by all causes that occurred within 30 days from the date of admission of the index hospitalization for hypertensive emergency. We defined 30-day readmission as hospitalizations for all causes occurring within 30 days from the date of discharge from the index hospitalization, using November 30, 2019, as the final date of discharge for complete follow up.^10-12^ We further defined the condition-specific readmission rates for the principal discharge diagnoses of congestive heart failure (CHF), acute myocardial infarction (AMI), and stroke (see **Supplemental eTable 1** for a list of ICD-9 and ICD-10 codes). If a patient had more than one hospitalization for hypertensive emergencies during the study period, we counted them all for the hospitalization analysis but randomly selected one for the mortality and readmission analysis.

### Statistical Analysis

We expressed the rates of hypertensive emergency hospitalizations as per 100,000 beneficiary-years, the rates of 30-day all-cause mortality and readmission as percentages. To assess trends in rates of hospitalization for hypertensive emergencies, we fit a mixed effects model with a Poisson link function and state-specific random intercepts, adjusting for age, sex, dual-eligible status, and race. To assess trends in the rates of 30-day mortality, we constructed a Cox proportional hazards regression to model the mortality as a function of patient age, sex, dual-eligible status, race, and comorbidity. We repeated this model for the 30-day readmission. Patients who switched to the Medicare Advantage plan after the initial hospitalization for hypertensive emergencies were treated as lost to follow-up, and deaths prior to a readmission were accounted using the Fine and Gray method^13^ for competing risks. We checked the adequacy of the Cox regression model and found that the proportional hazards assumption was satisfied.^14^ For all models, time was modeled as an ordinal variable, corresponding to the years 1999 (time=0) to 2019 (time=20), to represent the adjusted annual percent change in each outcome. We repeated models for age, sex, race, and dual status subgroups.

To assess geographic trends and variation in hospitalizations, we extended the CMS model used for profiling hospital performance on outcomes^13^ with a Poisson link function to model the number of hospitalizations for hypertensive emergencies as a function of patients’ age, sex, dual-eligible status, and race. We then calculated a risk-standardized number of hospitalizations per 100,000 beneficiary-years for each county and the beginning of (1999-2001) and ending (2017-2019) study periods. We mapped counties using a gradient from red, yellow, and green (increase in red, on average in yellow, and decrease in green).^15,16^ We also calculated a Pearson correlation coefficient between the 1999-2001 and 2017-2019 periods to evaluate the change in hospitalizations geographically.

All analyses were conducted using SAS version 9.4 (SAS Institute Inc.). All statistical testing was 2-sided at a significance level of 0.05. Institutional review board approval for this study was obtained through the Yale University Human Investigation Committee. The study followed the guidelines for cohort studies, described in the Strengthening the Reporting of Observational Studies in Epidemiology (STROBE) Statement: guidelines for reporting observational studies.^17^

## RESULTS

### Patient Characteristics and Comorbidities

The study sample consisted of 449,865 hypertension emergency-specific discharges (i.e., malignant hypertension, hypertensive heart disease, hypertensive renal disease, hypertensive emergency, hypertensive urgency, hypertensive crisis), represented 397,238 unique beneficiaries 65 years of age or older with ≥1 month of enrollment in the Medicare fee-for-service plan during the 21-year study period. Between 1999-2001 and 2017-2019, the average age of patients increased slightly (76.7 years [SD: 7.4] vs 77.9 years [SD: 8.7]), the proportion of female patients declined from 74.9% to 68.7%, White patients decreased from 74.9% to 70.7%, and Black patients increased from 20.3%to 21.3%. Several comorbidities were more commonly coded in 2017-2019 (**Table 1**), including renal failure (10.7% in 1999-2001 vs. 38.6% in 2017-2019), respiratory failure (2.5% in 1999-2001 vs. 7.5% in 2017-2019), and diabetes mellitus (29.6% in 1999-2001 vs. 39.2% in 2017-2019). All p values were <0.001 for trend.

**Table 1.** Characteristics of Patients Hospitalized for Hypertensive emergencies, 1999 to 2019.

| Patient Characteristics | No. (%) of Patients |  |  |  |  |  |  |
| --- | --- | --- | --- | --- | --- | --- | --- |
|  | 1999-2001 | 2002-2004 | 2005-2007 | 2008-2010 | 2011-2013 | 2014-2016 | 2017-2019 |
| No. of patients | 38181 | 27425 | 39117 | 43858 | 55931 | 94633 | 98093 |
| **Demographics** |  |  |  |  |  |  |  |
| Age, mean (SD), years | 76.7 (7.4) | 77.1 (7.6) | 77.3 (7.7) | 77.7 (8.0) | 77.9 (8.3) | 77.5 (8.5) | 77.9 (8.7) |
| Female | 28582 (74.9) | 20501 (74.8) | 28479 (72.8) | 31706 (72.3) | 39824 (71.2) | 60932 (64.4) | 67399 (68.7) |
| White | 28594 (74.9) | 20025 (73.0) | 27833 (71.2) | 30684 (70.0) | 38657 (69.1) | 63564 (67.2) | 69340 (70.7) |
| Black | 7747 (20.3) | 5893 (21.5) | 9139 (23.4) | 10579 (24.1) | 13579 (24.3) | 23416 (24.7) | 20903 (21.3) |
| Other races | 1840 (4.8) | 1507 (5.5) | 2145 (5.5) | 2595 (5.9) | 3695 (6.6) | 7653 (8.1) | 7850 (8.0) |
| Dual eligible | 8655 (22.7) | 6586 (24.0) | 9556 (24.4) | 10854 (24.7) | 14416 (25.8) | 24847 (26.3) | 21915 (22.3) |
| **Cardiovascular risk factors and conditions** |  |  |  |  |  |  |  |
| Diabetes | 11315 (29.6) | 8607 (31.4) | 13182 (33.7) | 15935 (36.3) | 21860 (39.1) | 42829 (45.3) | 38444 (39.2) |
| Renal failure | 4071 (10.7) | 3298 (12.0) | 7775 (19.9) | 17103 (39.0) | 23708 (42.4) | 51056 (54.0) | 37852 (38.6) |
| Atherosclerotic disease | 14267 (37.4) | 10770 (39.3) | 15778 (40.3) | 17966 (41.0) | 22585 (40.4) | 36571 (38.6) | 32313 (32.9) |
| Unstable angina | 1912 (5.0) | 1178 (4.3) | 1517 (3.9) | 1345 (3.1) | 1475 (2.6) | 3199 (3.4) | 3223 (3.3) |
| Prior myocardial infarction | 1251 (3.3) | 897 (3.3) | 1393 (3.6) | 1756 (4.0) | 2198 (3.9) | 4984 (5.3) | 3516 (3.6) |
| Prior heart failure | 6350 (16.6) | 4679 (17.1) | 7326 (18.7) | 8547 (19.5) | 10542 (18.8) | 24928 (26.3) | 14719 (15.0) |
| Peripheral vascular disease | 4401 (11.5) | 3306 (12.1) | 5111 (13.1) | 5879 (13.4) | 7110 (12.7) | 13196 (13.9) | 9088 (9.3) |
| Prior stroke | 1352 (3.5) | 1004 (3.7) | 1294 (3.3) | 1428 (3.3) | 1814 (3.2) | 3023 (3.2) | 3122 (3.2) |
| Cerebral vascular disease other than stroke | 4734 (12.4) | 3407 (12.4) | 4649 (11.9) | 5150 (11.7) | 6915 (12.4) | 10185 (10.8) | 12509 (12.8) |
| **Other conditions** |  |  |  |  |  |  |  |
| Chronic obstructive pulmonary disease | 6295 (16.5) | 4782 (17.4) | 7426 (19.0) | 8193 (18.7) | 9997 (17.9) | 19563 (20.7) | 15120 (15.4) |
| Pneumonia | 2783 (7.3) | 2172 (7.9) | 3495 (8.9) | 4551 (10.4) | 6174 (11.0) | 15035 (15.9) | 8726 (8.9) |
| Respiratory failure | 955 (2.5) | 701 (2.6) | 1331 (3.4) | 2328 (5.3) | 3503 (6.3) | 11113 (11.7) | 7390 (7.5) |
| Liver disease | 841 (2.2) | 702 (2.6) | 1062 (2.7) | 1304 (3.0) | 1757 (3.1) | 4207 (4.4) | 3445 (3.5) |
| Cancer | 1948 (5.1) | 1488 (5.4) | 2179 (5.6) | 2534 (5.8) | 3138 (5.6) | 6012 (6.4) | 5729 (5.8) |
| Depression | 2765 (7.2) | 2405 (8.8) | 3480 (8.9) | 4012 (9.1) | 5216 (9.3) | 8428 (8.9) | 9330 (9.5) |
| Other major psychiatric disorders | 1116 (2.9) | 854 (3.1) | 1246 (3.2) | 1474 (3.4) | 2167 (3.9) | 3414 (3.6) | 3346 (3.4) |
| Trauma in past year | 2049 (5.4) | 1613 (5.9) | 2595 (6.6) | 2933 (6.7) | 3663 (6.5) | 6227 (6.6) | 5878 (6.0) |

### Trends in Hospitalization for Hypertensive Emergencies Overall and by Demographic Subgroups

Between 1999-2019, the observed rate of hospitalization for hypertension emergencies among Medicare fee-for-service beneficiaries increased significantly from 51.5 to 125.9 per 100,000 beneficiary-years (**Figure 1**). Among Medicare beneficiaries, the increase in hospitalization for hypertensive emergencies was most pronounced among those who were ≥85 years of age (66.8 to 274.1), females (64.9 to 160.1), Black people (144.4 to 369.5), and dual-eligible (93.1 to 270.0; **Figure 2**). Across all age, sex, race, and dual-eligible strata, differences by race in hospitalizations for hypertensive emergencies increased substantially, with a rate of 369.5 per 100,000 beneficiary-years among Black beneficiaries compared with 104.8 per 100,000 beneficiary-years among White beneficiaries in 2019. These findings did not change substantially after adjusting for beneficiaries’ demographic characteristics. After adjusting for age, sex, race and dual-eligible status, the overall annual hypertensive emergency-hospitalization rate increased by 5.6% (95% CI 5.51-5.61); among Black beneficiaries the annual increase of 6.0% (95% CI 4.89-6.11) was more pronounced than in White beneficiaries (5.4%, CI 5.35-5.48) (**Supplemental eFigure 1**).

**Figure 1.**
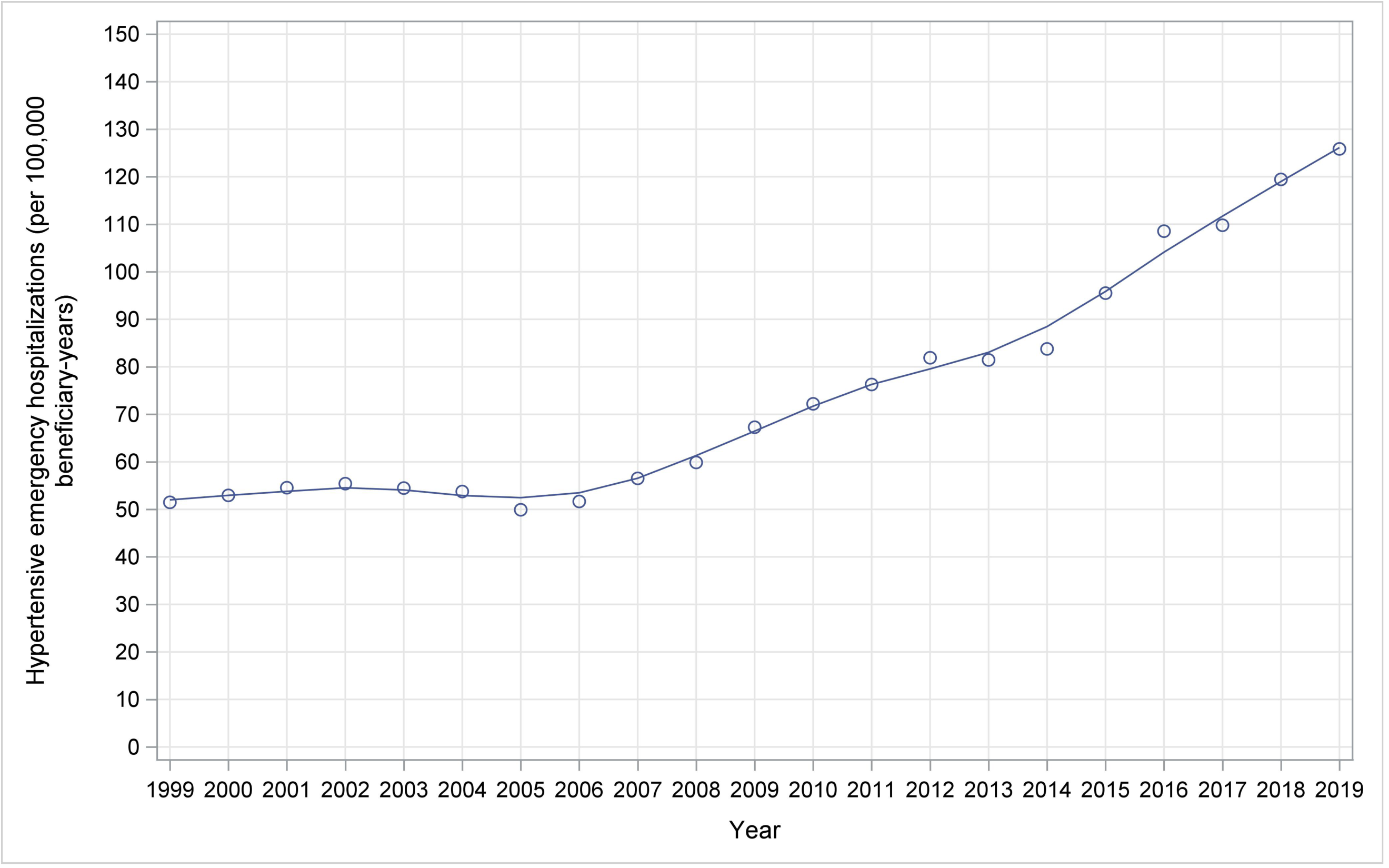
Observed Trends in Hospitalization Rate for Hypertensive Emergencies in the Medicare Fee-for-service Population, 1999-2019.

**Figure 2.**
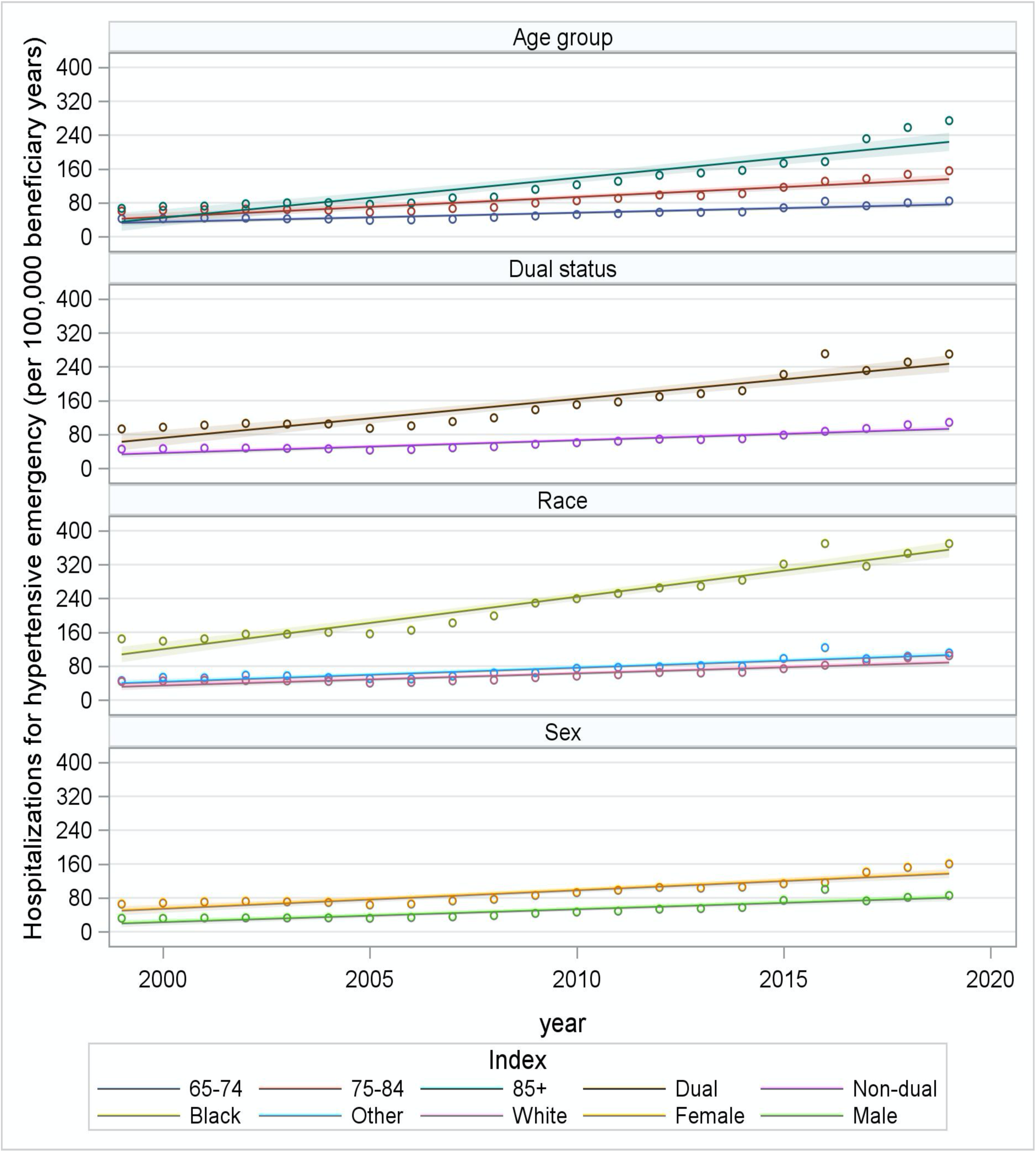
Trends in Observed Hospitalization Rate for Hypertensive Emergencies in the Medicare Fee-for-service Population by Age, Sex, Race, Dual Status, 1999-2019.

### Geographical Variation in Hospitalization for Hypertensive Emergencies

We observed marked geographic variation in hospitalization rates for hypertensive emergencies at the county level, with higher rates in the South, Mid-Atlantic, and Northwest in 1999-2001 (**Figure 3, panel A**); this was persistent in 2017-2019 (**Figure 3, panel B**). The Pearson correlation coefficient of county-specific hospitalization rates between 1999-2001 and 2017-2019 was 0.34 (95% CI: 0.30–0.37), indicating that the geographic pattern of hospitalization rates persisted moderately. The increase in hospitalization rates over the study period was observed geographically. Among 3,143 counties and county-equivalents included in the study, less than 1% counties either had no change (n=7) or decreased (n=20) hospitalization rates and the majority of counties (n=3,116) had increased hospitalization rates (**Figure 3, panel C**).

**Figure 3.**
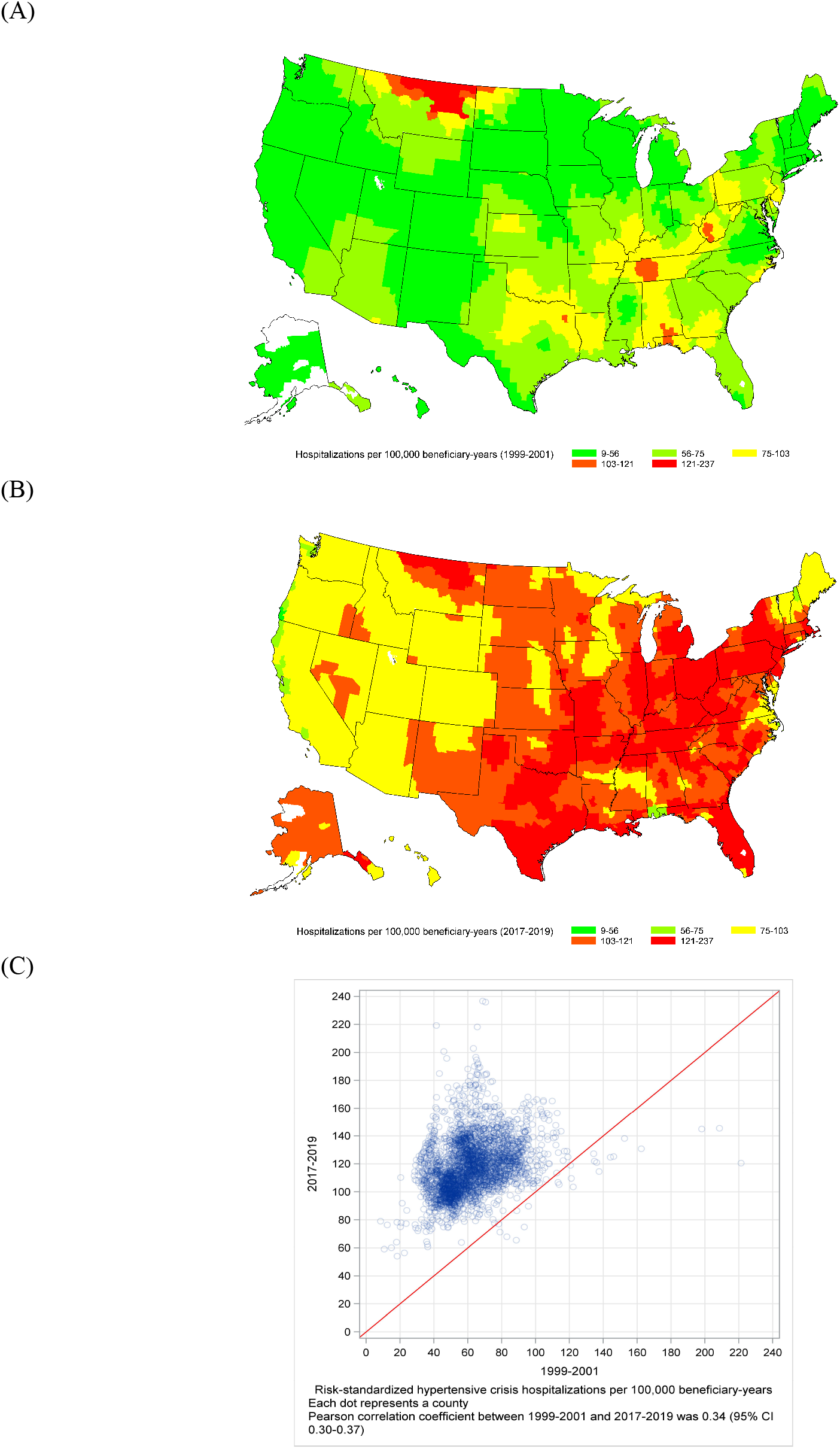
Maps Showing Trends in Risk-Standardized Hospitalization Rate for Hypertensive Emergencies in the Medicare Fee-for-service Population for Individual US Counties, 1999-2019. Panel A: Risk-Standardized Hospitalization Rate for Hypertensive Emergencies for Individual US Counties, 1999-2001. Panel B: Risk-Standardized Hospitalization Rate for Hypertensive Emergencies for Individual US Counties, 2017-2019. Panel C: Correlation Between Risk-Standardized Hospitalization Rate for Hypertensive Emergencies for Individual US Counties in 1999-2001 and 2017-2019.

### Trends in Mortality and Readmission Rates Among Hospitalized Patients

Among beneficiaries hospitalized for hypertensive emergencies, the observed 30-day all-cause mortality decreased from 2.6% (95% CI, 2.27-2.83) in 1999 to 1.7% in 2019 (95% CI, 1.53-1.80), and the observed 30-day all-cause readmission decreased from 15.7% (95% CI, 15.1-16.4) to 11.8% (95% CI, 11.5-12.1, **Supplemental eFigure 2**). No significant change was found in the rates of 30-day readmission for AMI (0.4% [95% CI 0.31-0.54] in 1999 and 0.4% [95% CI 0.30-.43] in 2019), but a marked decrease was observed in the rates of CHF from 2.5% (95% CI, 2.25-2.82) to 1.5% (95% CI, 1.42-1.68), in the rates of stroke from 0.8% (95% CI, 0.62-0.93) to 0.5% (95% CI, 0.46-0.61), and in the rates of AMI/CHF/stroke combined from 5.2% (95% CI, 4.78-5.58) to 3.8% (95% CI, 3.57-3.97).

These trends were similar among all age, sex, race, and dual status strata (**Figure 4**). After accounting for patient characteristics, the adjusted annual decrease rates were 2.3% (95% CI 2.20-2.45), 2.6% (95% ci 2.45-2.72), 2.5% (95% CI 2.35-2.61), 2.4% (95% CI 2.25-2.51), 2.4% (2.26-2.52), and 2.5% (2.37-2.64), for 30-day all-cause mortality, all-cause readmissions, CHF-specific, AMI-specific, stroke-specific, and AMI/CHF/stroke readmissions, respectively.

**Figure 4.**
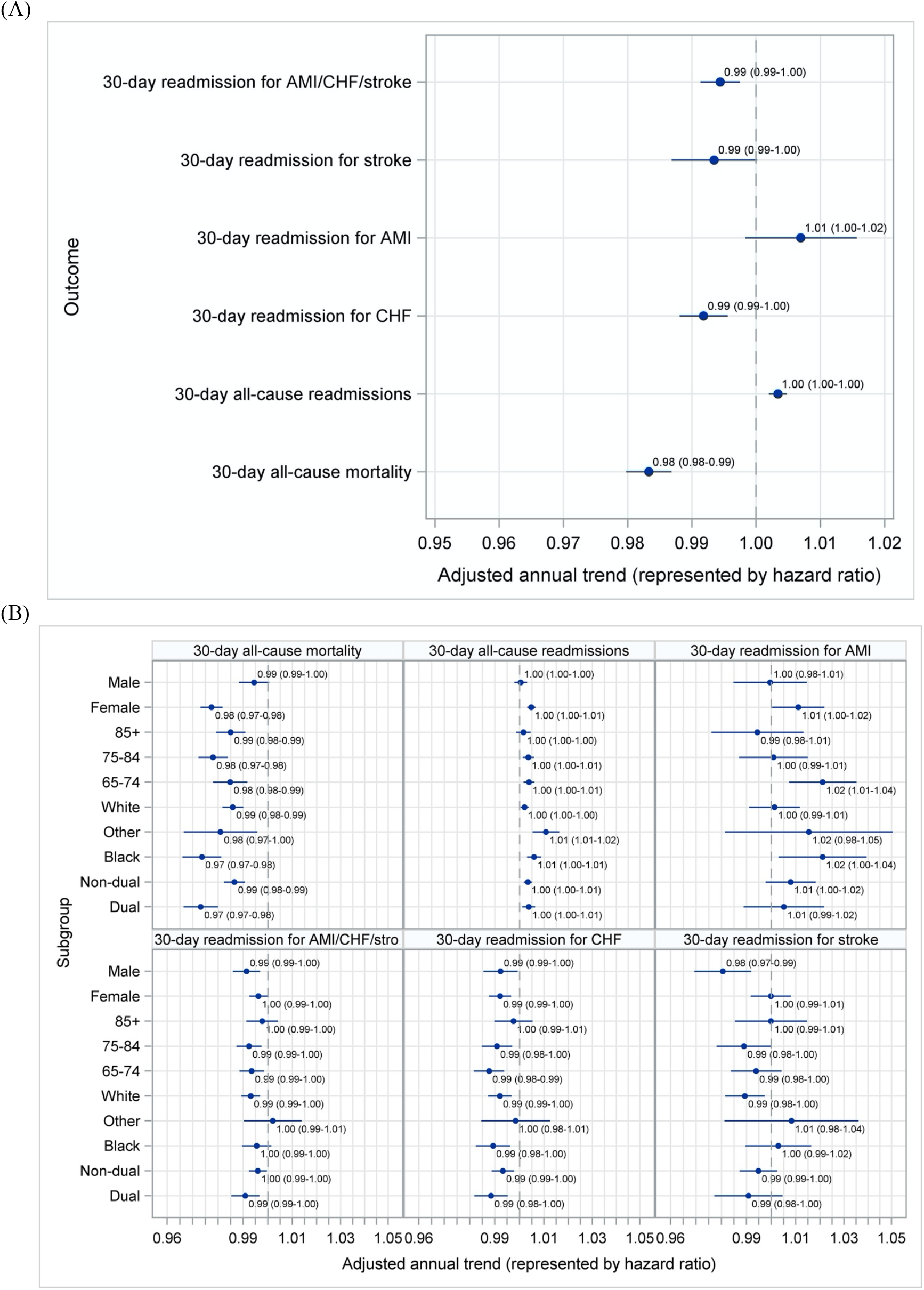
Average Annual Change in Hypertension Outcomes, Overall and by Subgroups. Panel A: Overall Average Annual Change in Hypertension Outcomes. Panel B: Overall Average Annual Change in Hypertension Outcomes by Subgroups.

## DISCUSSION

In this comprehensive analysis of the hospital trends in the Medicare fee-for-service population aged 65 years or older, we found a marked increase in hospitalization for hypertensive emergencies between 1999 and 2019. The increase was most pronounced among Black Medicare beneficiaries, the age group >85 years, women, and dual-eligible, with Black individuals having the largest 20-year increase across all age, sex, race, and dual-eligible strata. This overall trend was consistent nationwide, although geographic variation was present. Counties in the South (so-called “Stroke Belt”) had the highest rates of hospitalization.

This study adds to the literature in several ways. This study is the most contemporary study of the Medicare Fee-For-Service population and provides direct evidence of the failure of the clinical and public health interventions in reducing the risk of hospitalizations for hypertensive emergencies. The findings are concordant with evidence that age-adjusted mortality from hypertension is rising, stroke rates are rising, and hypertension control is declining. Moreover, Black adults have the highest rates of hospitalization for hypertensive emergency with the steepest increase in rates. Previous studies,^4-6^ with different analyses that used National Inpatient Sample data from 2000 to 2012, have showed results that were broadly consistent even as they focused on shorter time frames and did not examine variations by population subgroups.

Since the study uses administrative codes, it is important to examine if this may be artifactual. Polgreen and colleagues^6^ hypothesized that a shift towards assigning more severe diagnoses in administrative billing codes might have accounted for the increased hospitalization for hypertensive emergencies. However, the Polgreen’s study used data before 2011 and we also observed a marked increase in hospitalization for hypertensive emergencies rate after 2011. It seems implausible that coding shifts could have accounted for the marked changes over the whole 20-year period we observed. The increase in hospitalization was also unlikely associated with the change in ICD-9 to ICD-10 diagnosis coding in 2015 since the observed increase in hospitalization started in 2007. Moreover, the strongest case for the finding reflecting the true experience of patients is that we find consistency with the studies of hypertensive heart disease mortality, stroke mortality, and hypertension control. During the study period, hypertensive disease mortality rate among US adults aged 65 years and older increased from 98.4 in 1999 to 140.0 per 100,000 population in 2018.^18,19^ Stroke-related mortality rates declined from 1999 to 2011, and then markedly increased from 2012 to 2018 such that the stroke related mortality rate in 2018 was equal to the rate observed in 2000. Hypertension control rate among US adults aged 65 years and older decreased from 54.1% in 2013 to 44.5% in 2018.^1^ This consistent evidence suggests that the marked increase in hospitalization for hypertensive emergency, especially while health care costs are rising, are of great concern.

A striking finding of this study was highlighted by the rapid increase in racial disparities of hospitalization for hypertensive emergencies between Black and White beneficiaries, with Black individuals consistently having the higher rates during the last 20 years. Moreover, stratification by age, sex, and dual-eligible status showed persistent racial differences across strata. Such findings are especially important given hypertension disproportionally affects Black people. Compared with White people, hypertension in Black people occurs at an earlier age, is more severe, and is more likely to be associated with end-organ damage such as stroke, myocardial infarction, renal failure and death.^20,21^ Although there have been national attention and a wide variety of public health programs to address racial differences in health,^22-24^ this study demonstrated worsening health inequities with respect to acute hospitalization for hypertensive emergencies, highlighting the need for new approaches to address both medical and nonmedical factors that contribute to such inequities, including systemic racism,^25-29^ poor hypertension control,^30-32^ and high prevalence of renal diseases^33,34^ among Black individuals.

Furthermore, our study evaluates several longer-term outcomes after hospitalization for hypertensive emergencies that have not been studied in a national cohort. We showed improved mortality and readmission rates among those hospitalized and these improvements were consistent across age, sex, race, and dual-eligible subgroups. These improvements may, in part, be associated with national efforts to improve the care of all patients across the study period.^35-40^ Studies have showed improvements in process measures and outcomes for many conditions in Medicare beneficiaries.^15,16,41^ Additionally, improvements in outcomes observed may have been related to advance in treatments and health technologies. For example, the use of statins for prevention, the expansion of coronary revascularization, and the improvement in use of evidence-based medications are likely contributing to a lower risk of repeat hospitalization and improved survival associated with cardiovascular diseases.^42-44^ These results suggest that we have made progress in improving outcomes once patients are hospitalized for an acute illness; the issue is more about preventing hospitalizations for hypertensive emergencies.

### Limitations

This study has several limitations. First, we studied only Medicare fee-for-service beneficiaries and the results in this elderly population may not be fully generalizable to the broader US national population. However, this database represents the most complete national data for assessing rates of hospitalization and long-term outcomes. Second, we used administrative claims alone for the identification of hypertensive emergencies and could not confirm the diagnosis with blood pressure measurements and other clinically relevant prognostic factors. It is possible that changes in ICD coding practices could have affected our results. Nevertheless, previous studies by our group have shown that the performance of administrative data-based models for heart failure and myocardial infarction is comparable to that of the medical chart abstract-based models.^10,11^ Third, we defined hospitalization for hypertension emergencies based on primary discharge diagnosis and this will likely lead to underestimation of the number of hospitalization because some patients may have secondary diagnosis for hypertension emergency. Fourth, this study only examined hospitalizations and did not include Emergencies Department visits and outpatient visits. As a substantial proportion of the patients with hypertensive emergencies were managed in the Emergencies Department visits,^4-6^ our study is likely to underestimate the total medical needs of hypertensive emergencies. Fifth, we could not show what accounted for the changes in 30-day outcomes after hospitalization for hypertensive emergencies due to limited data available in the administrative claim database. Finally, we did not examine trends in Hispanic and other racial/ethnic subgroups due to misclassification of race in the Medicare data.^8,9^

## Conclusion

Among Medicare fee-for-service beneficiaries aged 65 years or older, hospitalization rates for hypertensive emergencies increased significantly from 1999 to 2019. Black beneficiaries had the largest increase in hospitalization rates for hypertensive emergencies across all age, sex, race, and dual-eligible strata. There was significant national variation, with the highest hospitalization rates generally in the South.

## Data Availability

Dr. Lu and Mr. Liu had full access to all the data in the study and take responsibility for the integrity of the data and the accuracy of the data analysis.

