## Supplemental material for "National Trends and Disparities in Hospitalization for Hypertensive Emergencies Among Medicare Beneficiaries, 1999–2019"

**eFigure 2.** Trends in Observed 30-day Mortality and Readmission Rates in the Medicare Fee-for-service Population, 1999-2019.

**eTable 1.** International Classification of Disease, Version 9 (ICD-9) and Version 10 (ICD-10) Codes Used for Hypertension Emergency, Acute Myocardial Infarction, Heart Failure and Stroke Identification.

**eTable 11.** Adjusted Annual Prevalence of No Health Care Utilization in the Past Year by Race and Latino/Hispanic Ethnicity, 1999-2018.

**eTable 12.** Adjusted Annual Prevalence of Foregone or Delayed Medical Care Due to Cost by Race and Latino/Hispanic Ethnicity, 1999-2018.

**eTable 13.** Adjusted Trends in the Prevalence and Racial/Ethnic Differences of Health Care Access, Utilization, and Affordability Measures by Race and Latino/Hispanic Ethnicity, Using Autoregressive Model, 1999–2018.

**eFigure 1**. **Average Annual Change in Hospitalization Rate for Hypertensive Emergencies, Adjusted for Age, Sex, Race, and Dual-eligible Status**

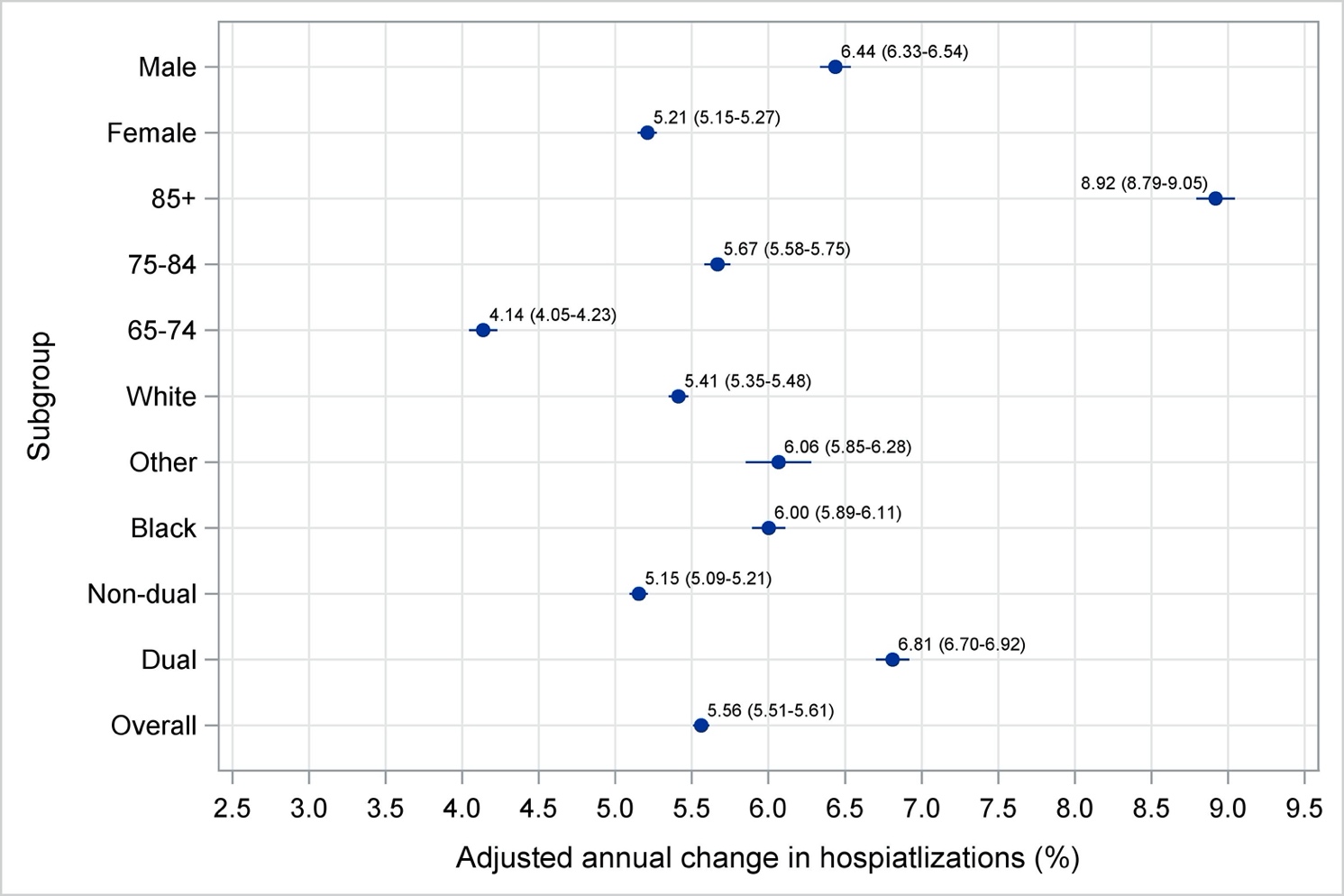

**eFigure 2. Trends in Observed 30-day Mortality and Readmission Rates in the Medicare Fee-for-service Population, 1999-2019**

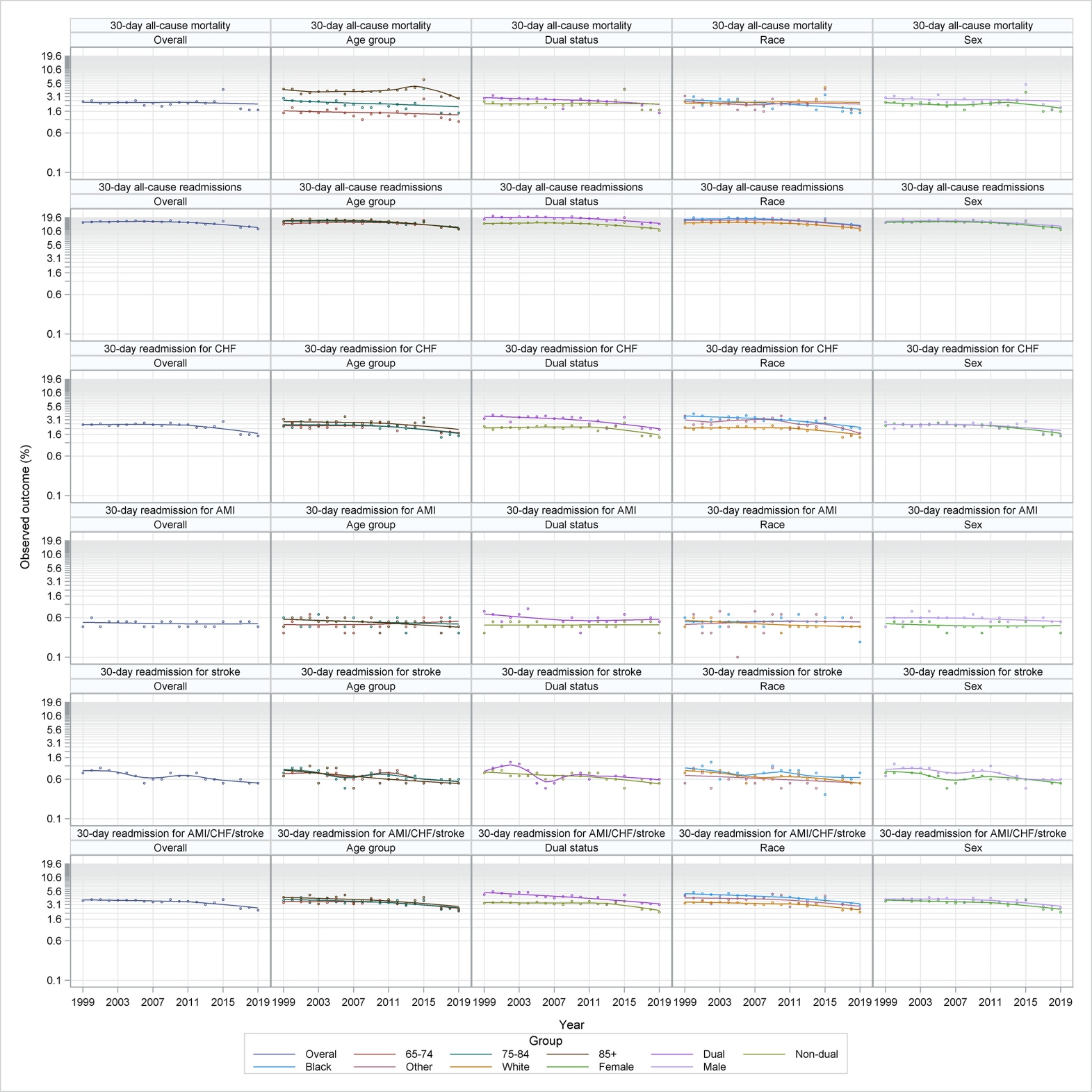

**eTable 1. International Classification of Disease, Version 9 (ICD-9) and Version 10 (ICD-10) Codes Used for Hypertension Emergency, Acute Myocardial Infarction, Heart Failure and Stroke Identification**

| **Condition** | **ICD-9 codes** | **ICD-10 codes** |
| --- | --- | --- |
| Hypertension Emergency or Hypertension Crisis | - 401.0 (malignant hypertension) - 402.0x (malignant hypertensive heart disease) - 403.0x (malignant hypertensive renal disease) - 404.0x (malignant hypertensive heart and renal disease) - 405.0x (malignant secondary hypertension) | From October 1, 2015, to September 31, 2016:   - I100 (hypertensive heart disease with heart failure) - I110 (hypertensive heart disease with heart failure) - I119 (hypertensive heart disease without heart failure) - I120 (hypertensive chronic kidney disease with stage 5 chronic kidney disease or end stage renal disease) - I132 (hypertensive heart and chronic kidney disease with heart failure and with stage 5 chronic kidney disease, or end stage renal disease) - I150 (renovascular hypertension) - I158 (other secondary hypertension)   From October 1, 2016, to December 31, 2019:   - I16.0 (hypertensive urgency) - I16.1 (hypertensive emergency) - I16.9 (hypertensive crisis) |
| Acute Myocardial Infarction | 410.xx, except 410.x2 | I21.0x I21.1x I21.2x I21.3 I21.4 I21.9 |
| Ischemic Stroke | 433.xx, 434.xx, 436.xx | I63.x |
| Heart Failure | 428.xx, 40201, 40211, 40291, 40401, 40411, 40491, 40403, 40413, 40493 | I50.21 I50.23 I50.31 I50.33 I50.41 I50.43 I50.811 I50.813 |
